# Velocity Reflection Index and Chronological Age: Confounder-Adjusted Statistical and Machine-Learning Analyses of Carotid Doppler Waveforms

**DOI:** 10.64898/2026.08.22.26361087

**Authors:** Azran Azhim

## Abstract

**Purpose:** To determine whether the velocity reflection index (VRI) is the carotid Doppler waveform feature most strongly associated with chronological age after adjustment for sex and exercise habit, and whether its feature ranking remains stable across cross-validated and cohort-sensitivity analyses.

**Methods:** Eight waveform-derived features were analysed in 197 participants meeting the study eligibility criteria and measured using a validated continuous-wave carotid Doppler system. Pearson and partial correlations and multivariable regression evaluated associations with chronological age. Random Forest regression with repeated 10-fold cross-validation, held-out permutation importance and bootstrap resampling assessed feature ranking. Sensitivity analysis evaluated the influence of cohort construction.

**Results:** VRI showed the strongest association with chronological age (r = 0.738, 95% CI [0.667, 0.796]) and remained strongly associated after adjustment for sex and exercise habit (partial r = 0.798). VRI ranked first by both impurity-based (0.536) and held-out permutation (0.765) importance; repeated cross-validation yielded MAE = 6.87 ± 1.26 years and R^2^ = 0.572 ± 0.153. Its leading ranking was stable in 85.3% of bootstrap resamples and the age-VRI correlation was essentially unchanged in the cohort-sensitivity analysis. The exercise association was significant after age adjustment (B = -0.043, p = 0.018) but attenuated after additional adjustment for sex (B = -0.026, p = 0.098). The sex association remained significant after adjustment for age and height.

**Conclusion:** VRI was robustly associated with chronological age and retained the leading feature-importance ranking across adjusted statistical and cross-validated machine-learning analyses. Validation against an established arterial-stiffness measure in an independent cohort is required before VRI can be considered a clinical vascular-aging biomarker.

## 1. Introduction

Vascular aging is characterized by progressive arterial stiffness: pulse-wave velocity accelerates, reflected waves return earlier in the cardiac cycle, and systolic hemodynamic burden increases, contributing to cardiovascular disease risk [1], [2], [14], [15]. Carotid Doppler waveform analysis provides physiologically meaningful information on vascular resistance, elasticity, and wave reflection [3-8], and waveform-derived indices have been linked to vascular aging and exercise adaptation [4-10]. The secondary systolic (reflected-wave) component arises from impedance mismatches at arterial branch points; as stiffness rises with age, the reflected wave returns earlier and merges with the incident wave during systole, producing an augmented secondary systolic peak (S2) and foreshortened incisura, the landmarks used to compute VRI here [24], [25]. Reference-standard tools (carotid-femoral pulse wave velocity, augmentation index, carotid intima-media thickness) require specialized equipment [15], [24]; Doppler-based analysis instead uses widely available, portable instrumentation [17], [18], not yet cross-validated against these measures in this cohort (Section 4).

Machine learning is increasingly applied to cardiovascular risk assessment [11], [12], but physiological interpretation of model outputs often remains limited [12], [13]. The present study evaluates VRI as a physiologically interpretable reflected-wave feature. Based on the wave-reflection mechanism above, we hypothesized that VRI would show a stronger and more consistent association with chronological age than the other seven waveform-derived features (S1, S2, I, D, d-min, RI and VEI). We tested this hypothesis using correlation and multivariable analyses together with cross-validated machine-learning feature importance, while identifying where validation against reference-standard vascular measures remains necessary.

Three related conference papers used eligible-participant subsets from the same source dataset to examine binary age classification [27], continuous age regression [28], and unadjusted associations of VRI with exercise, sex and body composition [29]. The present study provides an expanded archival analysis addressing a distinct question: whether the association between VRI and chronological age remains robust after confounder adjustment and alternative statistical and machine-learning evaluations. The additional contributions comprise joint adjustment for sex and exercise habit, held-out cross-validated permutation importance, bootstrap assessment of feature-ranking stability, incremental-value analysis, height-adjusted evaluation of the sex-VRI association, and direct sensitivity analysis of cohort construction. Previously reported findings are cited and interpreted as confirmatory context rather than new discoveries.

## 2. Materials and Methods

### 2.1 Subjects

Participants meeting the study screening criteria, aged 20-67 years, were enrolled (n = 197; 123 male, 74 female; per-decade distribution in Table 1); screening followed our group’s earlier reference-data study on this platform [3]: normotensive (systolic BP ≤ 140 mmHg), BMI ≤ 30 kg/m^2^, no cardiovascular, antihypertensive, or lipid-lowering medication, and no documented chronic cardiovascular disease. Screening relied on self-report and resting blood pressure, not laboratory-confirmed panels (Section 4). This dataset was first reported in Azhim et al. [3] for waveform-envelope reference data.

**Table 1.** Subject characteristics of the analysis set (n = 197), by age decade.

| Decade | n (total) | n male | n female |
| --- | --- | --- | --- |
| 20s | 92 | 56 | 36 |
| 30s | 36 | 15 | 21 |
| 40s | 20 | 12 | 8 |
| 50s | 36 | 30 | 6 |
| 60s | 13 | 10 | 3 |
| Total | 197 | 123 | 74 |

Overall: mean age $35.6 \pm 14.6$ years (range 20-67); 61 of 197 participants (31.0%) reported a regular exercise habit.

Participant flow: the source file contained 233 records. Thirty participants did not meet the medication-related eligibility criterion, and six records were missing all eight core waveform-derived features (d-min, S1, S2, I, D, RI, VRI and VEI). Exclusion of both groups yielded the final analysis cohort of 197 participants. For cohort-sensitivity analysis only, the originally compiled complete-waveform file comprised 227 records (233 source records minus the six waveform-incomplete records) before the medication-related re-screening. The age range narrowed from 20-89 to 20-67 years because all 30 medication-ineligible participants were aged 57 years or older. Figure 1 summarizes cohort construction and the analytical workflow; Table 2 compares the final analysis cohort with the 227-record sensitivity file.

**Table 2.** Final analysis cohort (n = 197) versus the originally compiled file used for sensitivity analysis (n = 227).

| Metric | Final analysis cohort<br>($n = 197$) | Sensitivity file<br>($n = 227$) |
| --- | --- | --- |
| Male / female | 123 / 74 | 136 / 91 |
| Age range (years) | 20-67 | 20-89 |
| Mean age ± SD (years) | $35.6 \pm 14.6$ | $40.2 \pm 18.3$ |
| Age-VRI correlation ($r$) | 0.738 | 0.739 |
| RF impurity-based importance (VRI) | 0.536 | 0.504 |
| 10-fold CV MAE (years) | $6.87 \pm 1.26$ | $8.27 \pm 1.40$ |
| 10-fold CV $R^2$ | $0.572 \pm 0.153$ | $0.609 \pm 0.140$ |

**Figure 1.**
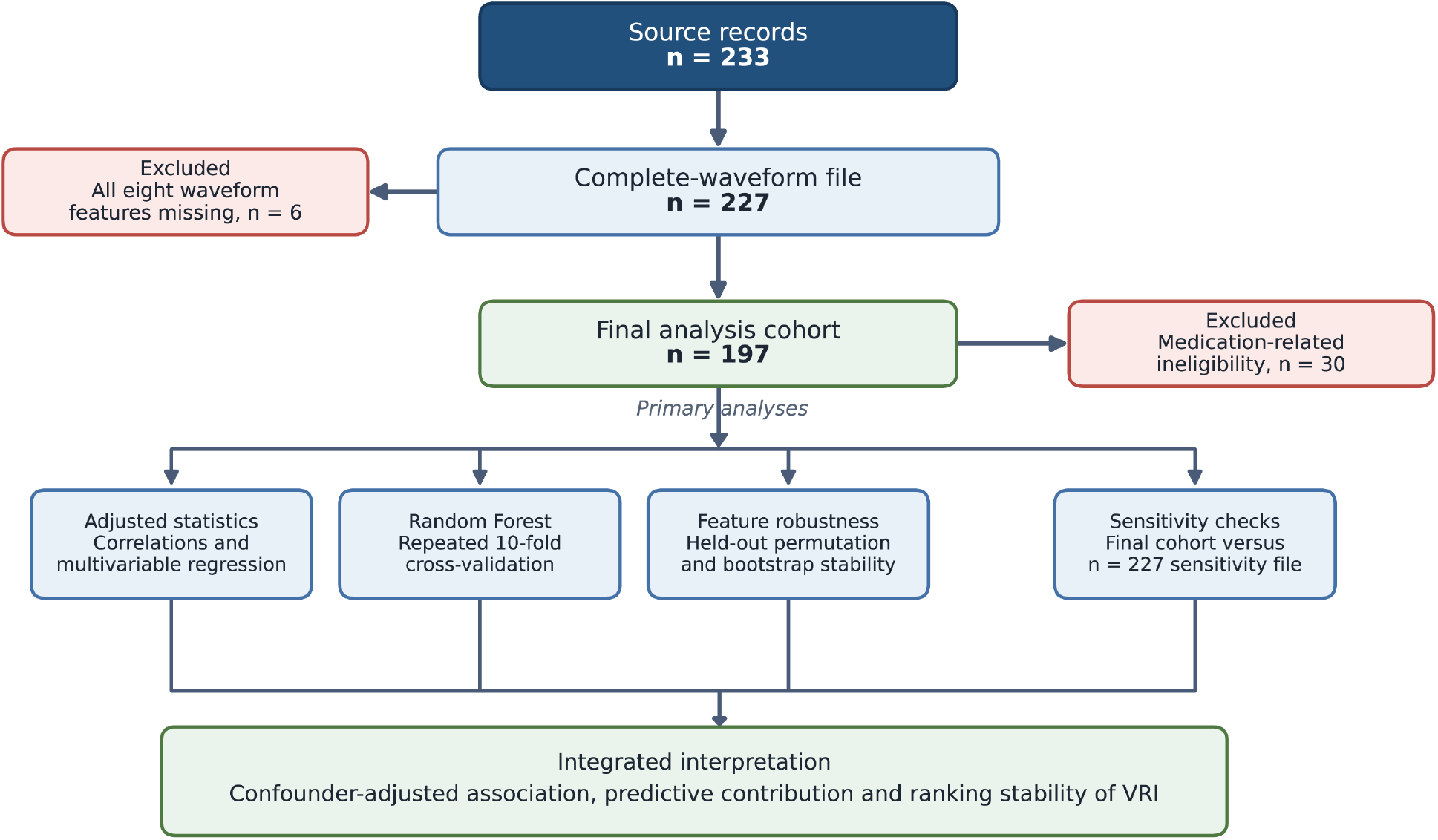
Participant selection and analytical workflow. The 227-record complete-waveform file was used only for cohort-sensitivity analysis; all primary analyses used the final 197-participant cohort.

The three companion studies used eligible-participant subsets: [27] used a balanced 82-participant classification subset (41 younger and 41 older), whereas [28] and [29] used a 202-participant subset that included 196 participants with complete eight-feature data. The present analysis includes 197 participants with complete eight-feature data. The one-record difference results from the wider age range used here: [28] and [29] were restricted to ages 20-65 years, whereas the present analysis extends to age 67 and includes one additional 67-year-old participant with complete data.

Overall: mean age 35.6 ± 14.6 years (range 20-67); 61 of 197 participants (31.0%) reported a regular exercise habit.

### 2.2 Doppler Blood Flow Velocity Measurement

Carotid waveforms were measured non-invasively using the custom wireless, portable continuous-wave Doppler system validated in [3]: a lightweight probe (two 15-mm piezoelectric transducers) on the left common carotid artery at a 50-degree insonation angle, 2.0 MHz irradiated frequency, with the demodulated signal band-pass filtered (0.1-4.2 kHz), digitized at 10 kHz, and telemetered for spectrogram analysis. The system was previously validated against a commercial ultrasonograph (r = 0.93, p < 0.05; mean S1-velocity difference 1.2 ± 0.5%) [3]. Following [3], the spectrogram was converted to a velocity envelope by threshold detection, and 30 artifact-free cycles were ensemble-averaged per participant.

### 2.3 Hemodynamic Biomarker Extraction

Biomarkers were extracted using the characteristic-point definitions and formulas established in [3], consistent with the Pourcelot/Gosling Doppler framework: five landmarks per cycle – peak systolic velocity (S1), second/reflected-wave systolic velocity (S2), incisura velocity (I), peak diastolic velocity (D), and end-diastolic minimum velocity (d). S1 reflects forward-traveling flow from ventricular ejection; S2 is primarily influenced by reflected waves from peripheral sites. Three dimensionless indices were computed:

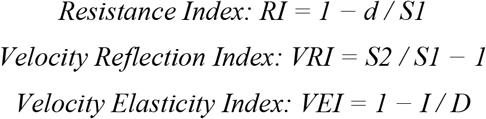

RI quantifies peripheral vascular resistance; VRI quantifies the relative magnitude of the reflected systolic component (normally negative; values closer to zero or positive indicate a larger relative reflected-wave component); VEI expresses proportional velocity decline between incisura and peak diastole, a surrogate for elastic recoil -- all three angle-independent ratios. In an earlier, overlapping cohort, partial correlations of RI, VRI, and VEI with age (adjusted for sex/exercise) were r = -0.57, 0.78, and -0.51 (all p < 0.001) [3], confirmed here in the final analysis cohort (n = 197) (partial r = -0.61, 0.80, -0.48; all p < 0.001; Section 4).

### 2.4 Statistical and Machine-Learning Analyses

Pearson correlation quantified linear associations between chronological age and the eight waveform-derived features (Figure 2); 95% confidence intervals used the Fisher z-transformation. Group comparisons used the Mann-Whitney U test because of non-normal distributions and were corroborated using Welch’s t-test. Statistical significance was set at p < 0.05, with exact p-values reported except when p < 0.001. Regression coefficients (B) are unstandardized and reported with 95% confidence intervals.

**Figure 2.**
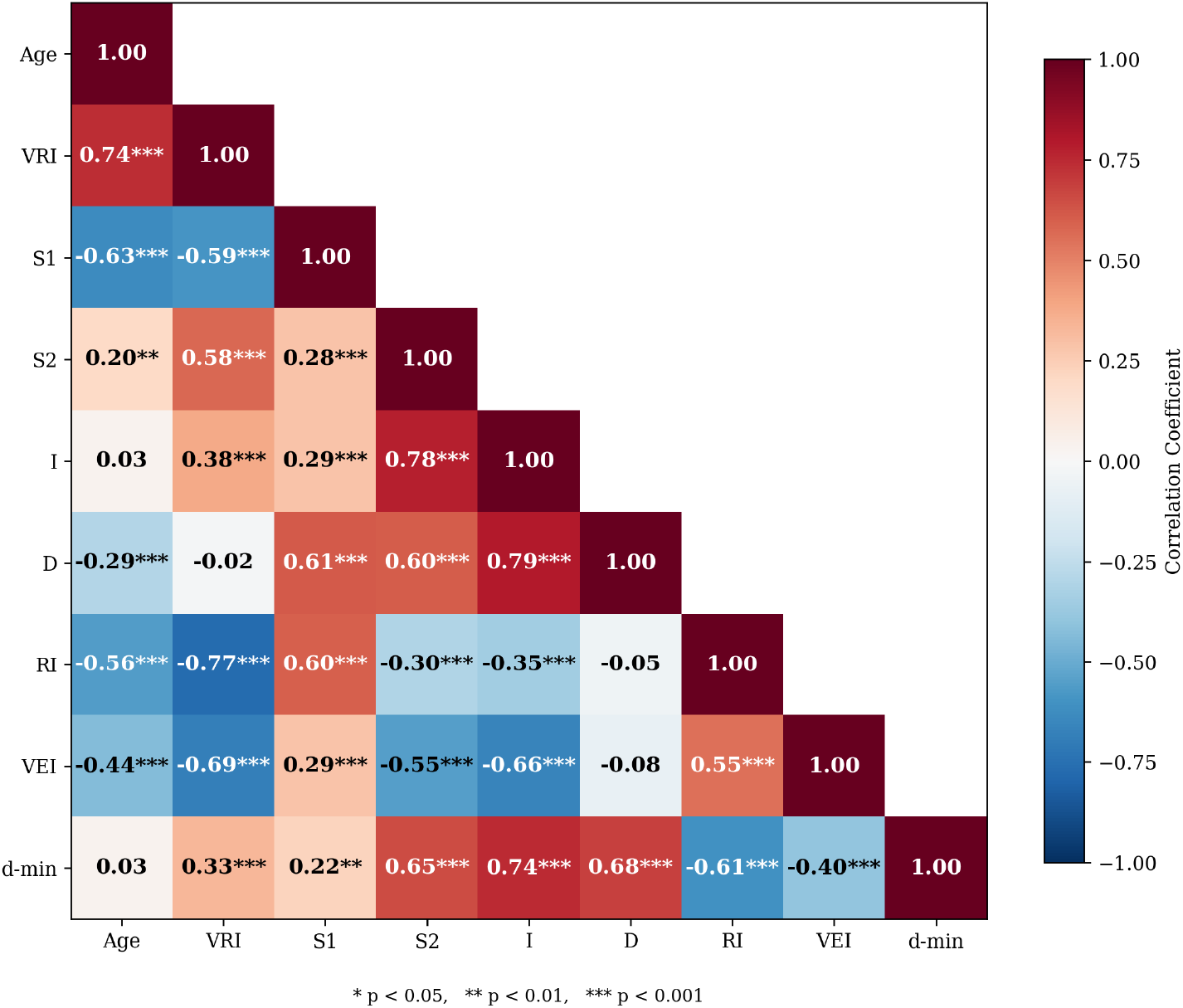
Correlation heatmap of the eight waveform-derived biomarkers with age.

Sex was coded 1 = male and 0 = female; exercise habit was coded 1 = yes and 0 = no. The age-adjusted exercise model used VRI ∼ age + exercise; the joint model used VRI ∼ age + sex + exercise; and the height-adjusted sex model used VRI ∼ sex + height + age. Interaction checks evaluated age by an indicator for age ≥60 years, sex or height. To evaluate whether VRI provided incremental information beyond the other waveform-derived features, two nested ordinary least squares regression models predicting chronological age were compared. The full model included all eight features (S1, S2, I, D, d-min, RI, VRI and VEI), whereas the reduced model included the same predictors except VRI. A partial F-test assessed the incremental contribution of VRI. Previously reported body-composition associations were not re-tested and are not presented as new findings.

A Random Forest regression model [11] predicted chronological age from the eight waveform-derived features using RandomForestRegressor (n_estimators = 500, max_features = None and random_state = 42; scikit-learn 1.8.0), with all other parameters retained at their software defaults. Performance was assessed by repeated 10-fold cross-validation (10 repeats; 100 test-fold estimates), yielding MAE, RMSE and R^2^. Impurity-based mean-decrease-in-impurity importance was used for initial feature ranking; the analysis was intended to assess feature contribution and ranking stability rather than clinical prediction. Because impurity-based importance can be biased when predictors are correlated, permutation importance was computed within held-out test folds using five repeats of 10-fold cross-validation (50 test-fold estimates), with each feature permuted 10 times per test fold. Bootstrap assessment of impurity-based ranking stability used 1,000 resamples. Benjamini-Hochberg correction was applied to the prespecified family of eight robustness tests. Analyses used Python with SciPy, pandas, scikit-learn and statsmodels.

## 3. Results

### 3.1 Correlation Analysis of Hemodynamic Biomarkers

In the final analysis cohort, age correlated most strongly and positively with VRI (r = 0.738, 95% CI [0.667, 0.796], p < 0.001) and negatively with S1 (r = -0.627), RI (r = -0.562), VEI (r = -0.435) and D (r = -0.290; all p < 0.001). S2 was weakly positively correlated with age (r = 0.201, p = 0.005), whereas I and d-min were not significant (r = 0.034, p = 0.630; r = 0.033, p = 0.650). These unadjusted results confirm the previously reported leading age association of VRI and provide the basis for the adjusted and robustness analyses below. Full 95% CIs are shown in Figure 2.

VRI also correlated with RI (r = -0.769), VEI (r = -0.686), and S1 (r = -0.586), with RI-VEI (r = 0.551) and VEI-S1 (r = 0.287) also correlated (all p < 0.001), indicating that reflected-wave intensity, elasticity, and systolic morphology are closely coupled -- relevant to the feature-importance ranking below (Section 3.2).

VRI showed the strongest overall age association, consistent with the study hypothesis; without mutual adjustment for exercise and sex here, or a gold-standard reference, this indicates increased wave reflection and reduced elasticity with vascular aging, not confirmatory validation of VRI (Section 4).

### 3.2 Cross-Validated Feature-Importance and Robustness Analysis

VRI had the highest impurity-based Random Forest importance (0.536; Figure 3), followed by S1 (0.218), I (0.058), VEI (0.052), S2 (0.045), D (0.035), RI (0.032) and d-min (0.025). Held-out permutation importance corroborated VRI’s leading ranking (mean decrease in R^2^ = 0.765 ± 0.178), approximately seven times the importance of S1 (0.110 ± 0.060), although the ordering of smaller features differed between methods. Because the leading VRI ranking was reported in the companion conference analyses [27]-[29], the present result is interpreted as a cross-validated robustness analysis rather than a new discovery. Bootstrap ranking stability and incremental-value analyses are reported in Section 3.5.

**Figure 3.**
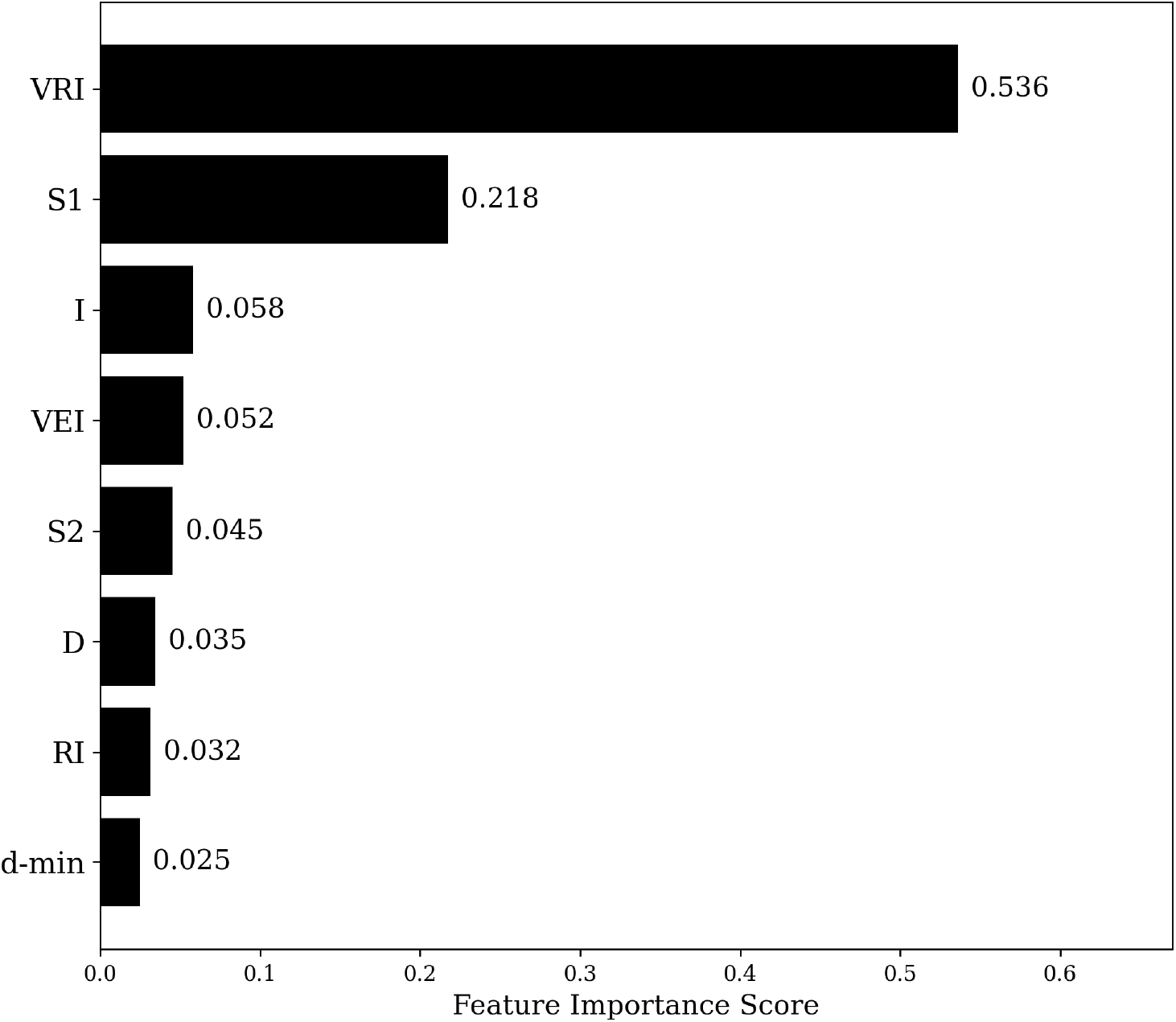
Random Forest impurity-based (Gini) feature importance for predicting chronological age from the eight waveform-derived biomarkers. This is a different metric from the held-out permutation importance reported in the text above; the two are not expected to yield numerically identical scores.

### 3.3 Exercise-Related Hemodynamic Adaptation

The unadjusted association between exercise habit and lower VRI was reported previously [29]. In the present cohort, the exercise group was younger than the non-exercise group (32.6 ± 13.6 versus 36.9 ± 14.8 years, p = 0.029). VRI remained lower with exercise after adjustment for age alone (B = -0.043, 95% CI [-0.078, -0.007], p = 0.018), but the association was attenuated and was not statistically significant after additional adjustment for sex (B = -0.026, 95% CI [-0.056, 0.005], p = 0.098; Figure 4). The cross-sectional and self-reported exposure does not support a causal exercise effect.

**Figure 4.**
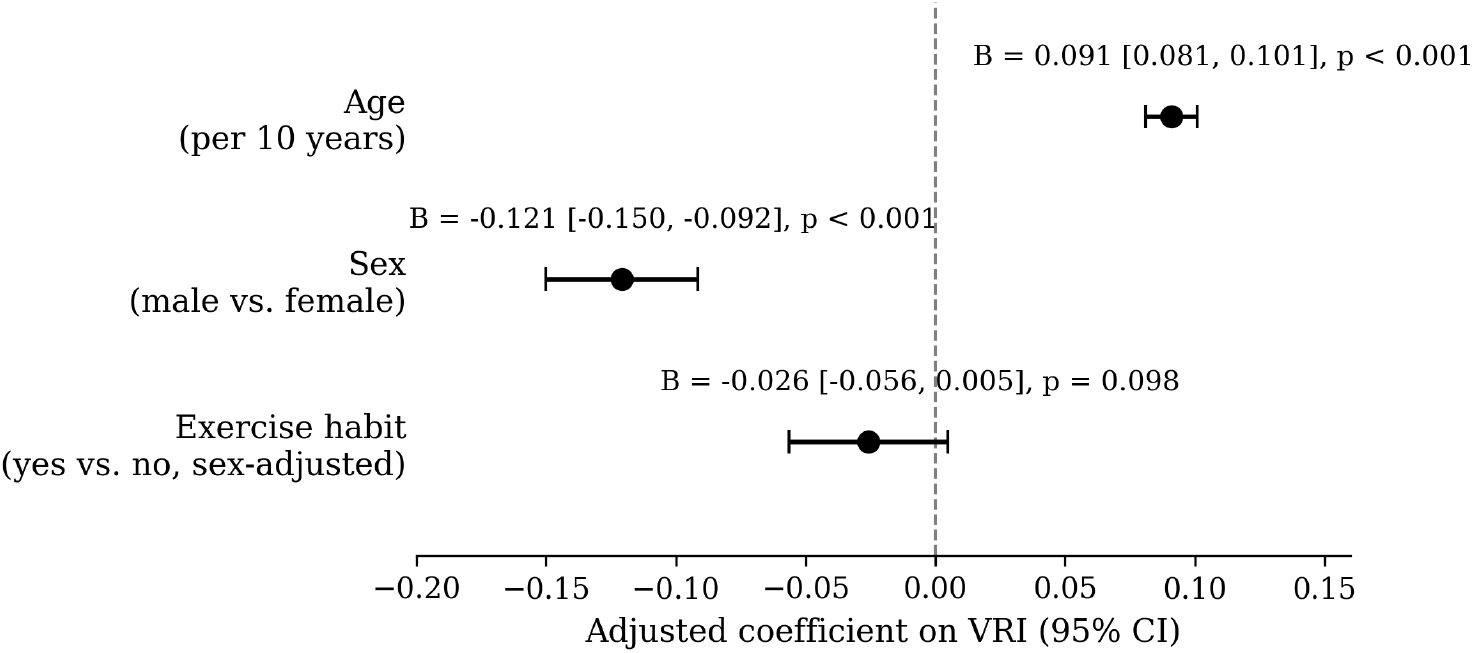
Confounder-adjusted association of age (per 10 years), sex (male vs. female), and exercise habit (yes vs. no) with VRI, from a multivariable regression (n = 197, R^2^ = 0.671); points are unstandardized regression coefficients (B) with 95% confidence intervals. The exercise coefficient here (B = -0.026, p = 0.098) is further sex-adjusted and smaller/non-significant, unlike the age-only estimate in Section 3.3 (B = -0.043, p = 0.018); the two are not directly comparable.

### 3.4 Sex-Dependent Wave Reflection Characteristics

The unadjusted VRI difference between male and female participants was reported previously [29]. The present analysis examined whether it was explained by body size. Among 145 participants with recorded height (89 male and 56 female), males were taller than females (169.8 ± 6.4 versus 157.5 ± 5.0 cm, p < 0.001), and height correlated negatively with VRI (r = -0.316, p < 0.001). Adjustment for height alone attenuated the sex-VRI association to non-significance (partial r = -0.075, p = 0.371). In a model including age and height (n = 145, R^2^ = 0.689), sex remained significant (p < 0.001), whereas height was borderline (p = 0.097), indicating that height did not fully account for the sex association. Previously reported unadjusted body-composition associations are not repeated because they were not subjected to a new multivariable analysis in the present study.

### 3.5 Sensitivity and Robustness Analyses

The originally compiled 227-record file was used only for sensitivity analysis. The age-VRI correlation and VRI’s impurity-based feature ranking were essentially unchanged, although cross-validated MAE and R^2^ differed because the final analysis cohort had a narrower age range. Restriction to age <60 years (n = 184) produced a similar correlation (r = 0.712). No age-by-older-group interaction was detected (p = 0.851, q = 0.907), but this analysis was underpowered because only 13 participants were aged 60 years or older. A quadratic age term added modest value beyond the linear model (R^2^ 0.545 to 0.565; p = 0.003, q = 0.008). Across 1,000 bootstrap resamples, VRI’s impurity-based importance averaged 0.516 (95% CI [0.213, 0.687]) and ranked first in 85.3% of resamples. Neither age-by-sex (p = 0.907, q = 0.907) nor age-by-height (p = 0.434, q = 0.578) interactions were detected. VRI added incremental value beyond the other seven features combined (R^2^ 0.593 to 0.646; partial F = 27.95, p < 0.001, q < 0.001), and VRI alone captured most of the full model’s explanatory power (R^2^ = 0.545 versus 0.646). Across the eight-test family, the age-VRI correlation, quadratic term, VRI increment and age-adjusted exercise association survived Benjamini-Hochberg correction; the interaction terms and height-adjusted sex-VRI partial correlation did not.

## 4. Discussion

VRI showed the strongest association with chronological age and retained the leading feature ranking across impurity-based, held-out permutation and bootstrap analyses. The confounder-adjusted and sensitivity results extend the previously reported unadjusted and Random Forest findings [27]-[29]. Because VRI integrates reflected-wave magnitude with systolic waveform morphology, its strong age association is physiologically plausible, but it does not establish equivalence with arterial stiffness or biological vascular age.

An important caveat applies throughout: this study lacked an established reference standard for vascular aging or arterial stiffness (e.g., carotid-femoral pulse wave velocity, augmentation index, carotid intima-media thickness) in the same subjects, and chronological age is an imperfect proxy for biological vascular aging. These results establish an association between VRI and chronological age and generate a hypothesis that VRI tracks arterial stiffness, rather than validating VRI as a clinical biomarker; direct comparison against a reference measure in an independent cohort is the most important next step.

The age-VRI association strengthened after adjustment for sex and exercise habit (partial r = 0.798 versus unadjusted r = 0.738), indicating that these variables did not explain the association. Exercise habit was associated with lower VRI after adjustment for age alone, but the estimate was attenuated and was not statistically significant after additional adjustment for sex. The sex association remained after adjustment for age and height, although body-size and body-composition relationships require further study in larger complete-case samples. The cross-sectional design does not establish causal direction for any association.

Held-out permutation importance provided a model-agnostic check of the impurity-based feature ranking and reduced reliance on a single Random Forest importance measure. Nevertheless, feature importance describes contribution to chronological-age prediction within this dataset and should not be interpreted as a causal explanation or as clinical biomarker validation.

### Limitations

The moderate sample was screened using self-report and resting blood pressure rather than laboratory-confirmed panels. The analysis was cross-sectional and lacked a reference-standard arterial-stiffness measure. Body-composition associations were not subjected to multivariable adjustment, and the data were acquired using a single portable Doppler device at one site. This cohort overlaps substantially with the eligible-participant subsets used in [27]-[29]; shared findings are treated as confirmatory context rather than new results. The final analysis cohort did not include adults older than 67 years, and only 13 participants were aged 60 years or older, limiting inference in older age groups. Independent validation incorporating a reference-standard measure and broader age distribution is required.

## 5. Conclusion

VRI was the carotid Doppler waveform feature most strongly associated with chronological age and remained strongly associated after adjustment for sex and exercise habit. Its leading feature-importance ranking was corroborated using held-out permutation importance, bootstrap resampling and cohort-sensitivity analyses. Previously reported exercise and sex associations were re-examined using adjusted models: the exercise association was attenuated after additional sex adjustment, whereas the sex association remained after adjustment for age and height. These results support VRI as an age-associated waveform feature, but validation against an established arterial-stiffness measure in an independent cohort is required before clinical application.

## Acknowledgements

The author acknowledges Tokushima University and the collaborators involved in acquiring and developing the carotid Doppler blood-flow-velocity measurements used in this study.

## Data availability

The de-identified data are not publicly available because of participant privacy and institutional restrictions. Requests may be considered subject to approval by the responsible institution and applicable ethics requirements. Analysis code is available from the corresponding author upon reasonable request.

## Declarations

### Ethics approval and consent to participate

Subjects were recruited through various forms of advertisement. All subjects provided written informed consent to participate. The study protocol and data collection were reviewed and approved by the Ethics Committee of Tokushima University Hospital, as previously reported in Azhim et al. [3]. The study was conducted in accordance with the Declaration of Helsinki.

### CRediT authorship contribution statement

A. Azhim: Conceptualization, Methodology, Software, Formal analysis, Investigation, Data curation, Writing – original draft, Writing – review & editing, Visualization.

